# De Novo Variation in Autism by Sex and Diagnostic Status in 41,367 Parent-Child Trios

**DOI:** 10.64898/2026.01.26.26344889

**Authors:** Tychele N. Turner

## Abstract

Autism shows a consistent sex bias, yet how sex shapes *de novo* variant (DNV) risk across coding and noncoding sequence remains unclear. We analyzed DNVs in 41,367 parent-child sequenced trios from three autism family-based cohorts and compared DNV characteristics and enrichment patterns in males and females. Importantly, these trios consisted of some trios with individuals with autism and some without autism. We developed a new sex-aware DNV caller and performed intensive, feature-based investigation of each candidate DNV to produce a high-confidence callset. We identified enrichment of missense and loss-of-function (LOF) DNVs both overall and within known autism-related genes (i.e., SFARI genes). Gene-specific enrichment analyses revealed twelve genes that were exome-wide significant and specific to males, for significance, including *FOXP1, SMAD6, AUTS2, CCDC168, PIEZO1, EML6, ZNF84, IGSF23, OTOG, SLC6A1, GIGYF1*, and *FREM3* and four genes that were specific to females, for significance, including *TAOK1, MECP2, DDX3X*, and *TBL1XR1* within a variant class. Direct comparisons of DNVs in males and females revealed *GABBR2* as the only gene trending toward enrichment in the direct males with autism comparison to females with autism. Finally, we analyzed promoters and identified a single significant promoter region (p = 3.8×10^-13^), associated with the *WDR74* gene, with the signal driven by DNVs observed in males with autism. Surprisingly, the noncoding RNA gene *RNU2-2* lies within this significant *WDR74* promoter and accounted for most of the DNVs in the region. *RNU2-2* DNVs were present in 0.2% of males with autism, and several are predicted to potentially alter RNA folding. We also observed *RNU2-2* DNVs in 0.2% of females with autism, including two DNVs that were recurrent (i.e., shared) with unrelated, affected males. Notably, *RNU2-2* DNVs were detected in 0.1% of unaffected males and were not observed in unaffected females. Together, these results suggest that although *RNU2-2* does not show a sex bias, it contributes to autism risk, which is intriguing due to a prior study implicating *RNU2-2* in a severe neurodevelopmental disorder.

## BACKGROUND

According to the Centers for Disease Control and Prevention, the overall prevalence of autism spectrum disorder is one in 31 individuals. Furthermore, ASD exhibits a sex bias with a prevalence in males of one in 20 individuals and in females of one in 70 individuals ^1^. Sex bias in human phenotypes and disease is common and the reasons for this bias are important for understanding of both the underlying mechanisms and ultimately for more tailored future clinical care. Potential mechanisms underlying sex bias include effects from the genome, hormones, and other potential environmental interactions. Regarding the genome, the most obvious culprits are the 1) sex chromosomes, themselves, and the loci which they interact with in the autosomal genome and 2) the genomic loci interacting with hormones. There are several potential downstream effects of this including gene expression differences in males and females. Classical examples of genes related to autism with effects from the X chromosome are *FMR1* repeat expansions in Fragile X Syndrome in males ^2^ and protein-affecting *de novo* variants (DNVs) in *MECP2* in Rett Syndrome ^3^. Fragile X syndrome preferentially affects males and Rett syndrome preferentially affects females due to differences in underlying mechanisms. An example of an effect from the Y chromosome is the significant increase in autism in individuals with XYY syndrome ^4^. A recent example indicating hormonal involvement in autism has shown a link between autism and fetal estrogens ^5^.

With the advent of whole-exome and whole-genome sequencing, the study of rare variants in autism genome-wide has expanded greatly. This has yielded several studies identifying an excess of rare and *de novo* variants in females with autism as compared to males with autism ^6, 7^. In autism, this phenomenon has taken on the identity of the female protective effect ^8^. More generally comparisons of rare variants have been expanded out beyond autism to a broader set of individuals including those with developmental disorders. This is important; however, it is also relevant to note that developmental disorders more broadly do not exhibit as substantial of a skewing towards males ^7^. The result of these studies has been the identification of several genes on the X chromosome with enrichment in females with developmental disorders with the most common being *DDX3X* ^*9*^. This gene is a gene on the X chromosome that escapes X inactivation and thereby is expressed at higher levels in females with autism than males with autism presumably providing a type of “buffered” expression.

Since the sex bias in autism is higher than in general developmental disorders, the focus of this current study is on DNVs in large cohorts of families with autism. The most concerted efforts to sequence individuals with autism has come from the Simons Foundation through their cohorts including the Simons Simplex Collection ^10^ and the Simons Powering Autism Research (SPARK ^11^). In this study, >41,000 parent-child trios, sequenced by either whole-exome sequencing (WES) or whole-genome sequencing (WGS), are assessed for DNVs in the protein-coding regions of the genome and a subset of >9,800 parent-child trios sequenced by WGS are assessed for DNVs in the noncoding regions of the genome.

In this study, we generate high-quality, sex-aware, DNVs using data from multiple sequencing technologies and characterize DNV patterns across three autism family cohorts. We evaluate gene-set and gene-specific enrichment of loss-of-function (LOF) and missense DNVs in males and females with or without autism, including analyses involving the autosomes, the pseudoautosomal regions (PARs), the non-PAR X chromosome, and the non-PAR Y chromosome. We also test for enrichment of DNVs in promoter regions in males and females with or without autism, respectively.

## METHODS

### Study Cohorts

Three cohorts consisting of families with autism were accessed from the Simons Foundation Autism Research Initiative (SFARI) Base. These included 4,173 parent-child sequenced trios with whole-genome sequencing (WGS) data from the Simons Simplex Collection (SSC) ^10^, 5,675 parent-child sequenced trios with WGS data from Simons Powering Autism Research (SPARK ^11^), and 32,487 parent-child sequenced trios with WES data from SPARK.

### De novo variant calling

For WGS data, family-level variant call files (VCFs) were obtained from a previous study ^12^. Briefly, these VCFs were generated from genotyping Genomic VCFs (gVCFs) generated by GATK HaplotypeCaller ^13^ and DeepVariant ^14^, respectively, with GLnexus ^15^. For WES data, we downloaded per-individual gVCFs generated with DeepVariant from SFARI Base (accession: SPARK iWES_v3). For each family, we performed joint-genotyping using GLnexus (release v1.4.1-0-g68e25e5).

For both WGS and WES, we called DNVs using HAT-FLEX version 1.0 (https://github.com/TNTurnerLab/HAT-FLEX). Briefly, HAT-FLEX is a Python post-calling computational tool for identifying DNVs from parent-child trios using one or two genome-wide VCFs per trio (e.g., GATK, DeepVariant), with an optional two-caller intersection mode; it supports human and other diploid species, reads trios from a simple family file or standard pedigree file, and applies sex-aware haploid/diploid logic on X/Y including awareness of PARs, so that filtering and parent relevance depend on genomic context. It detects candidate DNVs via sex-aware genotype patterns (reference alleles in parent(s), alternate allele in child), optionally splits multi-allelic sites by allele with normalization, can mask user-provided regions, and then applies layered quality controls including required FORMAT keys, strand-bias checks, depth/GQ thresholds, parent “ALT leak” rules (allele depth and genotype-based), allele-balance thresholds (diploid and haploid), homopolymer handling, and a “haploid unusual” flag for heterozygous-like signals on haploid loci. Outputs include a sorted, normalized trio-only VCF with added INFO/FILTER annotations (e.g., DENOVO, SEXCTX, IRRELPARENT, HAPUNUSUAL, CLUSTER), optional TSV and metrics/manifest summaries, and automatic bgzip compression plus optional tabix indexing (with gzip fallback), along with logging, and verbosity controls.

For WGS data, the parameters of HAT-FLEX included --ped to input a standard pedigree, --caller1_vcf for the DeepVariant family VCF, --caller2_vcf for the GATK HaplotypeCaller VCF, --intersect_mode of allele to intersect the two callers by allele, --normalize_alleles set to true, --par_bed set to build 38 coordinates, -- gq_value set to 20, --depth_value set to 10, --haploid_depth_value set to 5, --ab_min set to 0.25, -- haploid_ab_min set to 0.85, --require_format_keys set to GT,DP,AD,GQ, --strict_format set to false, -- max_homopolymer set to 10, --cluster_window_bp set to 100, and --cluster_min_count set to 2. For WES data, the same parameters were used except for --caller2_vcf and --intersect_mode, which were not used, since only DeepVariant was used in the HAT-FLEX calling on the WES data.

Post-calling of DNVs, we collapsed DNV calls by merging nearby child-only variants into single records when they met strict eligibility criteria. Variants were processed in genomic order per contig, requiring the child to carry an alternate allele while all other samples were homozygous reference, and restricting merging to high-confidence PASS sites annotated with the HAT-FLEX cluster flag. Eligible variants within 100 bp were grouped into clusters; for clusters with multiple variants, we defined the minimal spanning interval, retrieved the reference haplotype, and assembled the alternate haplotype by substituting the alternate allele at each site while disallowing overlaps. We required at least two reads to support the assembled (collapsed) haplotype. Next, we interrogated alignment data for evidence of the alternate allele at candidate DNVs by scanning parental and child reads around each variant annotated with the HAT-FLEX DENOVO flag. Using the child VCF and the child, father, and mother CRAM files, we counted ALT-supporting reads in each individual. Support was assessed at the haplotype level from aligned query-reference base pairs, with explicit handling of SNVs, MNVs, and INDELs; reads were required to anchor the reference position and, for INDELs, to match the expected inserted or deleted sequence. For variants annotated with SEXCTX=PAR, we also queried the homologous PAR on the partner sex chromosome to capture shared coverage. We required zero high-quality ALT-supporting reads in both parents and at least two high-quality ALT-supporting reads in the child. Finally, we inferred parent-of-origin for DNVs in trios by combining phase-by-transmission generated on a genome-wide trio VCF with read-backed physical phasing from the alignment data of the child. We first identified informative transmission sites genome wide as high-quality biallelic SNVs where the child was heterozygous and all trio genotypes had GQ ≥ 20, excluding non-PAR X/Y in male probands. For each DNV, we collected child reads supporting the ALT allele and phased them to nearby informative SNVs within ±40 kilobase pairs (kbp) by determining which parental allele those same reads carried. We summed paternal and maternal support across informative sites to assign parent-of-origin by majority vote (ties or insufficient evidence were reported as unknown), and quantified confidence using a binomial test reported as both a 0 to 1 score and a Phred-scaled value. For male probands, non-PAR X and Y calls were assigned deterministically (X assigned maternal, Y assigned paternal).

### Generating a Cross-Cohort DNV Callset

The DNVs called above were annotated for their presence in the Human Pangenome Reference Consortium (HPRC) ^16, 17^. First, the decomposed VCF from the HPRC was downloaded from https://s3-us-west-2.amazonaws.com/human-pangenomics/pangenomes/freeze/freeze1/minigraph-cactus/hprc-v1.1-mc-grch38/hprc-v1.1-mc-grch38.vcfbub.a100k.wave.vcf.gz. Each DNV was tested for pangenome-graph membership. Each DNV was queried against the pangenome VCF for presence or absence at the site and/or the allele level. DNVs present at the site-level in the pangenome VCF were removed from further analyses. QC of the DNVs was performed using acorn ^18^. For WGS data, individuals with <= 40 DNVs or > 150 DNVs were removed from downstream analyses (n = 24 individuals in SSC and n = 23 individuals in SPARK). For WES data, individuals with >8 DNVs were removed from downstream analyses (n = 40 individuals). DNVs were than annotated with Variant Effect Predictor (VEP) ^19^ for predicted functional consequences. DNVs that are SNVs were summarized as an SBS96 mutational spectrum using deconstructSigs ^20^. Single base substitution signatures were visualized by hierarchical clustering heatmaps (individuals × signatures) using heatmap.2 ^21^ with dendrograms for both rows and columns. For the any analyses involving exome data, to ensure comparable callability across samples, analyses were restricted to DNVs in the targeted capture regions.

### Gene Set Enrichment Testing

Missense and LOF DNVs were assessed for gene set enrichment using the denovolyzeByClass function in denovolyzeR ^22^. Our analyses evaluated enrichment across eight gene sets including 1) all genes, 2) non-PAR genes on the X chromosome, 3) non-PAR genes on the Y chromosome, 4) PAR genes, 5) hormone receptor genes from the GtoPdb ^23^ accessed at https://www.guidetopharmacology.org/GRAC/NHRListForward, 6) genes exhibiting sex-bias in gene expression in the brain ^24^, 7) genes present in the SFARI Gene database (release 2025 Q4) (https://gene.sfari.org/), and 8) 100 random genes exclusive of the SFARI gene list. For each gene set, enrichment was tested for synonymous DNVs, missense DNVs, and LOF DNVs.

We analyzed four DNV datasets stratified by proband sex (male/female) and affection status (affected/unaffected). After basic cleaning and filtering (including harmonizing gene symbols), we formatted each dataset for denovolyzeR and performed gene set enrichment tests. We considered a gene set significant if it passed Bonferroni correction for 24 tests (p < 0.002).

Since denovolyzeR does not stratify expected DNV counts by child sex, we adjusted expectations for non-PAR genes on the X and Y chromosomes using a parent-of-origin model with male:female germline mutation-rate ratio *r* = *μ*_male_/*μ*_female_ = 3 (i.e., paternal fraction = *r*/(1 + *r*) = 0.75). denovolyzeR’s baseline expectation is *E* ∝ 2*Np*_DNM_, where *N*is the number of probands and *p*_DNM_is the per-gene, per-class probability from denovolyzeR’s internal table; let *N* = *N*_*m*_ + *N*_*f*_ with *N*_*m*_males and *N*_*f*_ females. Relative to the autosomal baseline ∝ (*μ*_male_ + *μ*_female_), transmission implies for non-PAR loci: male X ∝ *μ*_female_ and male Y ∝ *μ*_male_, yielding multipliers *x*_male_ = 1/(1 + *r*) and *y*_male_ = *r*/(1 + *r*); female multipliers are *x*_female_ = 1.0 and *y*_female_ = 0.0. For mixed-sex groups, *x*_mult_ = (*N*_*f*_ ⋅ 1.0 + *N*_*m*_ ⋅ *x*_male_)/*N* and *y*_mult_ = (*N*_*m*_ ⋅ *y*_male_)/*N*. PAR genes were unadjusted (multiplier = 1.0). Implementation-wise, we multiplied the numeric columns of denovolyzeR’s probability table by gene-specific multipliers and supplied the adjusted table to denovolyzeByClass via the probTable argument; genes without a specified multiplier were left unchanged (multiplier = 1.0).

### Per-Gene Enrichment Testing

Per-gene DNV enrichment was tested separately within each sex × affection stratum using denovolyzeR (denovolyzeByGene) for LOF, missense, and LOF+missense classes, with non-PAR X/Y expectations adjusted as described above. Genes with a p-value less than 2.5 × 10^-6^ were considered exome-wide significant.

### Male Versus Female DNV Comparisons

As a complementary analysis, we tested per-gene enrichment using two-sided Fisher’s exact tests on allelic variant counts. For each gene and chromosomal category (autosomes, PAR, X non-PAR), we constructed four pairwise comparisons: (1) affected females vs affected males, (2) unaffected females vs unaffected males, (3) affected females vs unaffected females, and (4) affected males vs unaffected males. For each comparison, we formed a 2×2 table of variant alleles vs non-variant alleles between the two groups, where the group-specific denominator equaled the number of allele copies evaluated given cohort size and chromosome copy number (autosomes/PAR: 2N per group; X non-PAR: 2N in females and N in males). Non-PAR Y genes were not included in between-group comparisons. Genes with a p-value less than 2.5 × 10^-6^ were considered exome-wide significant.

### Testing for DNV Enrichment in Promoter Regions

Promoters were tested for enrichment of DNVs using fitDNM ^25, 26^. They were defined as 2000 bp upstream of the transcription start site of protein-coding genes from the RefSeq database downloaded from the UCSC Genome Browser ^27, 28^. Due to the discovery, in this study, of *RNU2-2*, in a genome-wide significant promoter we also tested for enrichment of DNVs in the gene based on coordinates from the UCSC Genome Browser.

### Modeling RNA Folding of RNU2-2 Wild Type and Alternate Sequences

We extracted the sequence for *RNU2-2* from the reference genome and converted it to transcript-oriented RNA (reverse-complementing negative-strand and replacing T with U). We then generated RNA sequences for each individual based on the *RNU2-2* haplotypes present in their genome. RNA secondary-structure minimum free energies (MFEs) were computed with ViennaRNA ^29^, and stability effects were summarized as ΔΔG=MFE_mut_−MFE_WT_. In parallel, we performed *in silico* single-nucleotide saturation mutagenesis across the wild-type sequence (all possible A/C/G/U substitutions at each position) to obtain a position-by-mutation ΔΔG landscape; results were summarized in tabular outputs and visualized with lollipop and density plots.

## RESULTS

### Characteristics of High Quality DNVs in this Study

Throughout the remainder of the paper the following acronyms are used to represent the cohorts. “SSC WGS” refers to whole-genome sequencing data from the Simons Simplex Collection, “SPARK WGS” refers to whole-genome sequencing data from the SPARK cohort, and “SPARK WES” refers to whole-exome sequencing data from the SPARK cohort. For the SSC WGS cohort, we identified 279,272 SNVs, 49,404 INDELS (16,658 deletions, 32,746 insertions), and 2,660 MNVs. The mean ± standard deviation of length for the deletions were 4.76 ± 5.42 bp, for insertions was 5.75 ± 5.86 bp, and for MNVs was 14.06 ± 20.71 bp. For the SPARK WGS cohort, we identified 394,476 SNVs, 55,182 INDELS (21,489 deletions, 33,693 insertions), and 3,284 MNVs. The mean ± standard deviation of length for the deletions were 5.15 ± 5.98 bp, for insertions was 5.95 ± 6.26 bp, and for MNVs was 12.55 ± 18.99 bp. For the SPARK WES cohort, we identified 49,516 SNVs, 5,384 INDELS (1,404 deletions, 3,980 insertions), and 384 MNVs. The mean ± standard deviation of length for the deletions were 7.31 ± 7.99 bp, for insertions was 6.63 ± 4.08 bp, and for MNVs was 11.69 ± 14.42 bp (**Figure 1A**).

**Figure 1.**
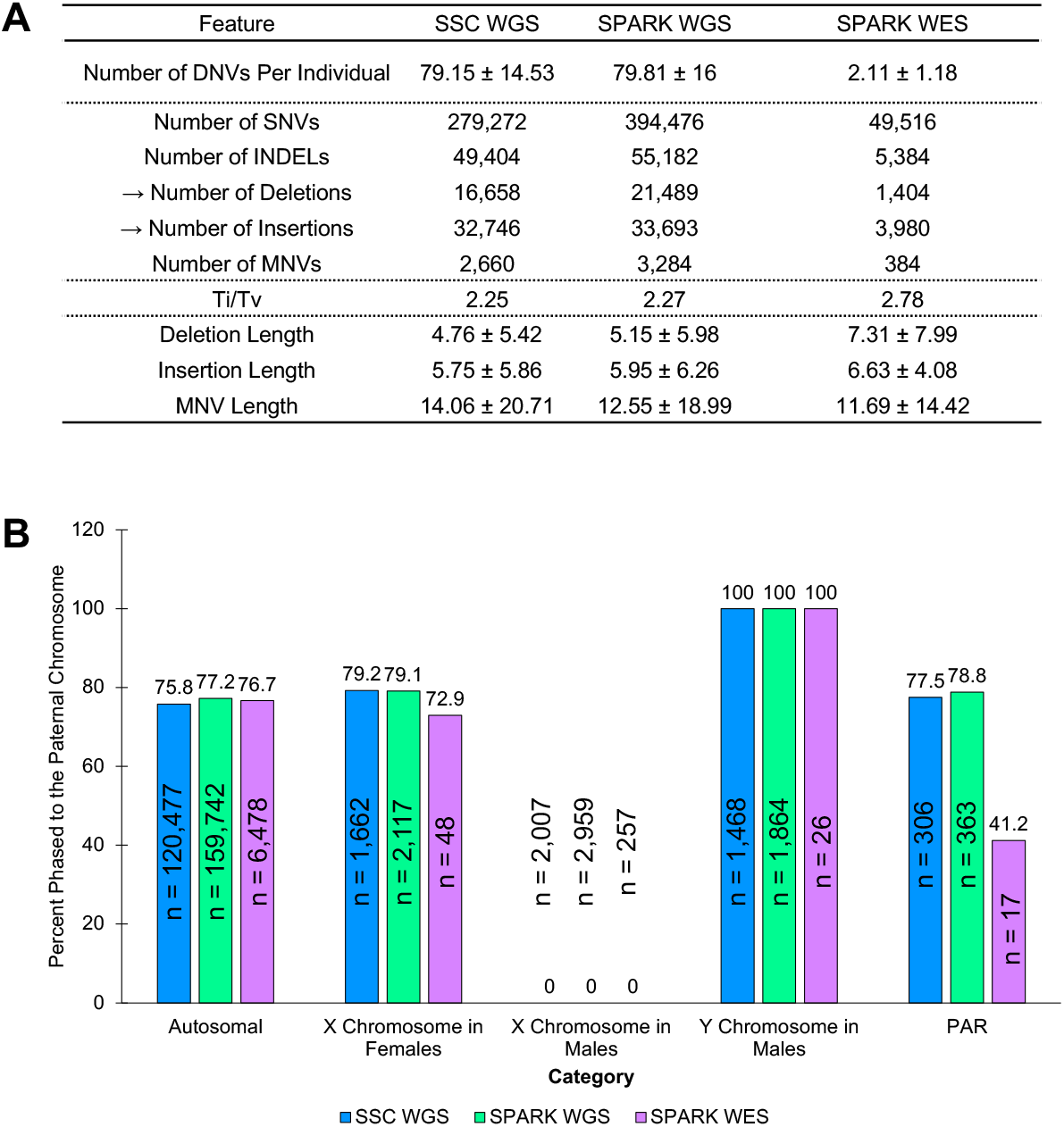
Overall Features of DNV Callset Generated in this Study. Three cohorts are shown including Simons Simplex Collection whole-genome sequencing (SSC WGS), Simons Powering Autism Research (SPARK) WGS, and SPARK whole-exome sequencing (SPARK WES). A) Shown are the number of DNVs per individual (expect ∼75 in WGS and ∼2 in WES), the number of variants by type, the Ti/Tv ratio (expect ∼2 in WGS and ∼2.8 in WES), and the mean ± standard deviation of length for various DNV types. B) Shown is information on DNVs that could be phased for parent-of-origin chromosome. Inside each bar is the count of DNVs, on the y-axis are the percent phased to the paternal chromosome, on the x-axis is the category (i.e., genomic region). For autosomal, X chromosome in females, and PAR, it is expected that ∼75% are phased to the paternal chromosome of origin. For the X chromosome in males, 0% are expected to be phased to the paternal chromosome. For the Y chromosome in males, 100% are expected to be phased to the paternal chromosome.

Several metrics were considered for determination of high quality DNVs (**Figure 1A**). First, the number of DNVs per individual was calculated and was 79.15 ± 14.53 in SSC WGS, 79.81 ± 16 in SPARK WGS, and 2.11 ± 1.18 in SPARK WES. Second the Ti/Tv ratio was calculated and was 2.25 in SSC WGS, 2.27 in SPARK WGS, and 2.78 in SPARK WES. Genome-wide the percent of DNVs that could be phased to the parent-of-origin chromosome was 38.0% in SSC WGS, 36.9% in SPARK WGS, and 12.3% in SPARK WES. Third, phased variants were considered in 5 different categories including the autosomes, the X chromosome in females, the X chromosome in males, the Y chromosome in males, and the pseudoautosomal regions (PAR). All DNVs in males on the X chromosome implicitly phase to the maternal chromosome and on the Y chromosome phase to the paternal chromosome (**Figure 1B**). For the autosomes, the percent phased to the paternal chromosome was 75.8%, 77.2%, and 76.7% in SSC WGS, SPARK WGS, and SPARK WES, respectively (**Figure 1B**). For the X chromosome in females, the percent phased to the paternal chromosome was 79.2%, 79.1%, and 72.9% in SSC WGS, SPARK WGS, and SPARK WES, respectively (**Figure 1B**). For the PARs, the percent phased to the paternal chromosome was 77.5%, 78.8%, and 41.2% in SSC WGS, SPARK WGS, and SPARK WES, respectively (**Figure 1B**). The lower percent in SPARK WES is likely due to a small DNV count of 17 that could be phased in that category.

WGS data were used to assess additional characteristics of the DNVs. Overall, there was an increase in DNV counts with parental age at birth (**Figure 2A, Figure 2B**). A linear model of DNV counts and paternal and maternal ages at birth, respectively, were assessed and identified an increase of 1.4 [1.3,1.5] DNVs per year for paternal age and 0.5 [0.4,0.6] DNVs per year for maternal age in SSC WGS with an adjusted r^2^ for the model of 0.45. For SPARK WGS, these values were an increase of 1.5 [1.4,1.6] DNVs per year for paternal age and 0.5 [0.4,0.6] DNVs per year for maternal age with an adjusted r^2^ for the model of 0.54. For the combined dataset, these values were an increase of 1.5 [1.4,1.5] DNVs per year for paternal age and 0.5 [0.4,0.5] DNVs per year for maternal age with an adjusted r for the model of 0.50. Overall, a shift towards older fathers compared to mothers in this study (**Figure 2C**).Next, single base substitution signature analyses were performed on the DNV data. The top contributing signatures were Signatures 1A, 1B, 5,and 16 (**Figure 2D**). This is consistent with prior findings in DNV datasets ^30^.

**Figure 2.**
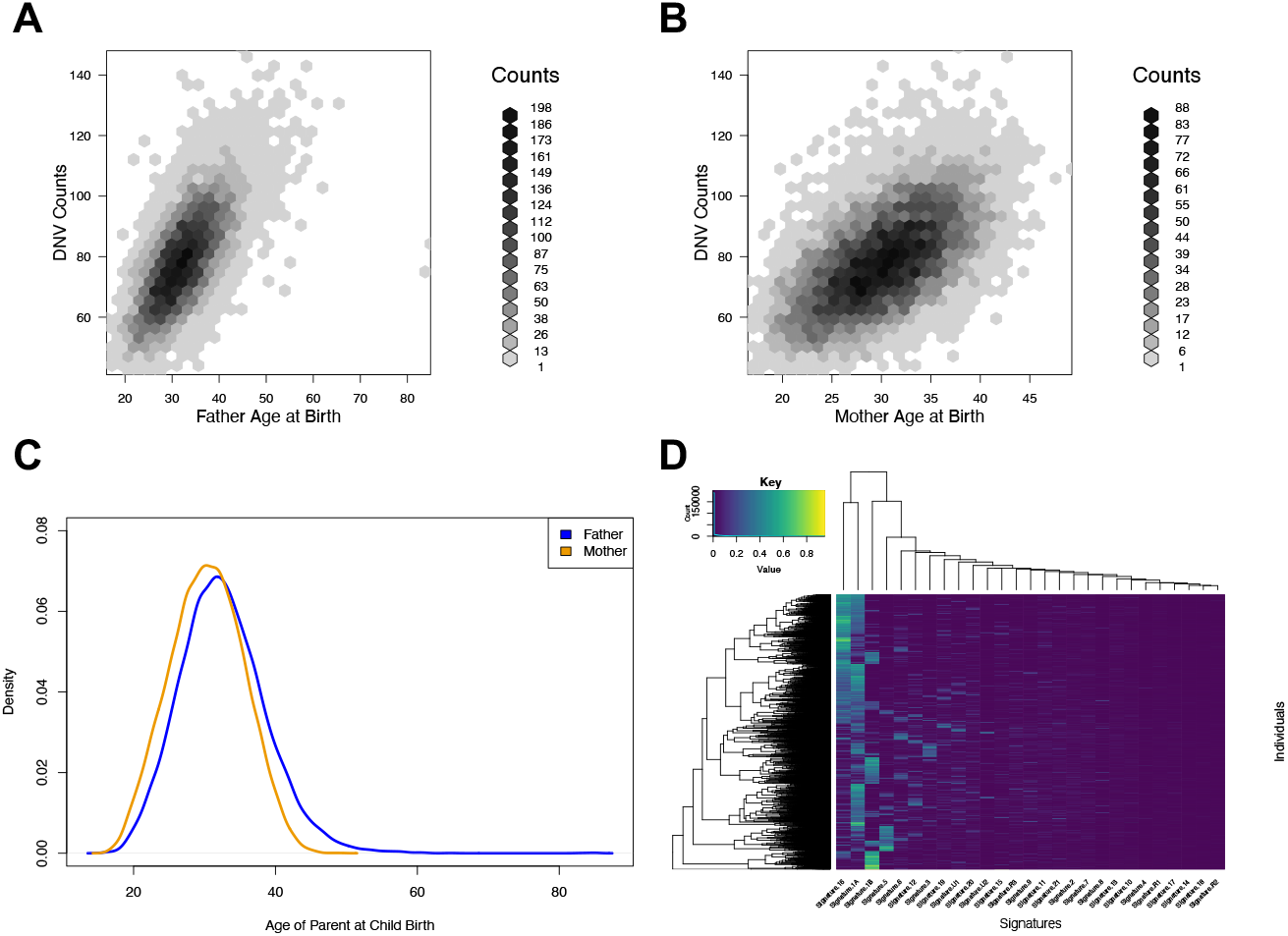
DNVs Increase with Parental Age at Birth and Show Expected Clock-Like Mutation Signatures. A) Hexagonal binning plot of DNV counts per individual versus the age of the father at birth. B) Hexagonal binning plot of DNV counts per individual versus the age of the mother at birth. C) Density plot showing the ages of father and mother at birth. Fathers in this study are, generally, slightly older than mothers. D) Heatmap of mutation signatures generated using deconstructSigs. Each row is an individual, and each column is a signature. The color corresponds to the fraction of that signature contributing to the individual signature profile. The top signatures are the expected clock-like signatures including Signature 1A, Signature 1B, Signature 5, and Signature 16.

### Gene-Based Enrichments Reveal Specific Categories

We performed gene set enrichment on the combined WGS data and the combined WGS and WES data, respectively. For both analyses, four different groups were assessed including males with autism (n = 4,810 individuals in WGS, n = 22,081 individuals in the combined WGS/WES set), females with autism (n = 1,024 individuals in WGS, n = 5,937 individuals in the combined WGS/WES set), males without autism (n = 1,928 individuals in WGS, n = 6,481 individuals in the combined WGS/WES set), and females without autism (n = 2,086 individuals in WGS, n = 6,868 individuals in the combined WGS/WES set). The testing in each group was across eight different gene sets (all genes, non-PAR X chromosome genes, non-PAR Y chromosome genes, PAR genes, hormone receptor genes, genes with sex-bias in expression in the brain, SFARI genes, and random genes) and three variant types (synonymous, missense, LOF).

In the WGS dataset (**Figure 3A**), no group had significance for synonymous variants in any category. In males with autism, there was a significant enrichment of missense DNVs in the all gene set (p = 7 × 10^-10^, enrichment = 1.11), the LOF DNVs in the all gene set (p = 4 × 10^-15^, enrichment = 1.41), missense DNVs in the SFARI gene set (p = 2 × 10^-39^, enrichment = 1.77), and LOF DNVs in the SFARI gene set (p = 6 × 10^-73^, enrichment = 4.55). In females with autism, there was a significant enrichment of missense DNVs in the all gene set (p = 3 × 10^-5^, enrichment = 1.16), LOF DNVs in the all gene set (p = 4 × 10^-11^, enrichment = 1.76), LOF DNVs in the non-PAR X chromosome genes (p = 3 × 10^-6^ , enrichment = 4.72), missense DNVs in SFARI genes (p = 1 × 10^-17^, enrichment = 2.1), and LOF DNVs in SFARI genes (p = 2 × 10^-47^, enrichment = 8.0). In unaffected males, there was an enrichment of missense DNVs in SFARI genes (p = 0.0004, enrichment = 1.3). In unaffected females, there was an enrichment of missense DNVs in the non-PAR X chromosome genes (p = 8 × 10^-8^, enrichment = 1.92).

**Figure 3.**
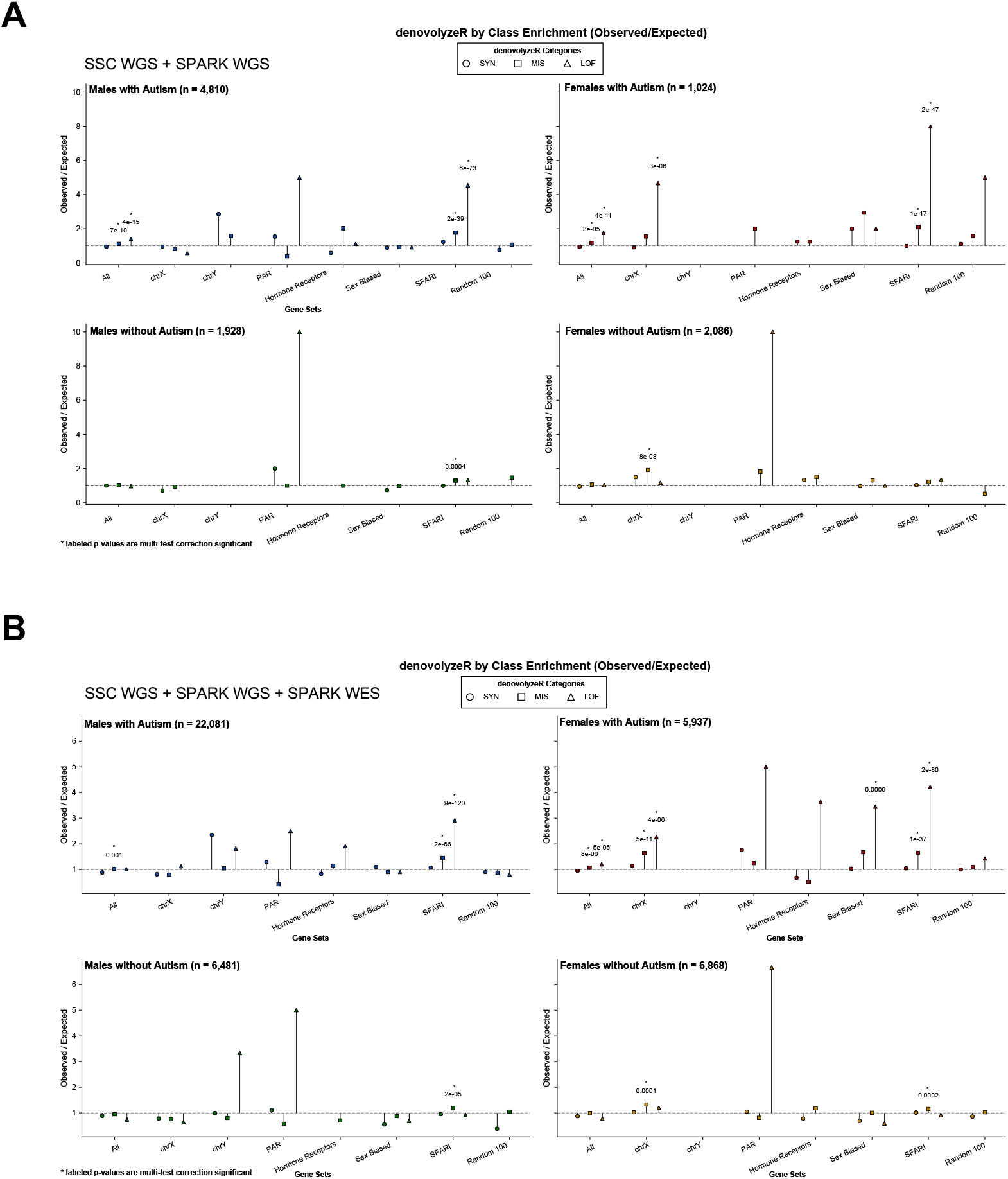
Gene Set Enrichment Testing. Shown are gene set enrichment analyses generated using denovolyzeR for four different settings (males with autism, females with autism, males without autism, females without autism). Eight different gene sets are considered in the analyses and within each gene set analysis three different variant types are considered including synonymous, missense and LOF. A) Results shown for the combined WGS data and B) results shown for the full combined WGS and WES data. Multi-test correction significant p-values are labeled and have a ^*^.

In the combined WGS/WES dataset (**Figure 3B**), no group had significance for synonymous variants in any category. In males with autism, there was a significant enrichment of missense DNVs in the all gene set (p = 0.001, enrichment = 1.03) and in the SFARI gene set (p = 2 × 10^-66^, enrichment = 1.45). There was also a significant enrichment of LOF DNVs in the SFARI gene set (p = 9 × 10^-120^, enrichment = 2.92). In females with autism, there was a significant enrichment of missense DNVs in the all gene set (p = 8 × 10^-6^, enrichment = 1.07), the non-PAR X chromosome genes (p = 5 × 10^-11^, enrichment = 1.65), and SFARI genes (p = 1 × 10^-37^, enrichment = 1.65). There was a significant enrichment of LOF DNVs in the all gene set (p = 5 × 10^-6^), enrichment = 1.20, the non-PAR X chromosome genes (p = 4 × 10^-6^, enrichment = 2.27), the genes with sex-bias in expression in the brain (p = 0.0009, enrichment = 3.45), and SFARI genes (p = 2 × 10^-80^, enrichment = 4.22). In unaffected males, there was an enrichment of missense DNVs in SFARI genes (p = 2 × 10^-8^, enrichment = 1.19). In unaffected females, there was an enrichment of missense DNVs in the non-PAR X chromosome genes (p = 0.0001, enrichment = 1.33) and the SFARI genes (p = 0.0002, enrichment = 1.16).

### Gene-Based Enrichment of DNVs

We first performed denovolyzeR analyses for LOF, missense, and combined LOF and missense DNVs in the full set of individuals with autism (n = 28,018 individuals). The 25 genes significant in the LOF category were *SCN2A, CHD8, ADNP, SHANK3, GRIN2B, PTEN, MIB1, CHD2, FOXP1, POGZ, SMAD6, MARK2, SYNGAP1, GIGYF1, BRSK2, KDM6B, KDM5B, DLG4, AUTS2, ASH1L, TANC2, ARHGEF9, CSNK2B, SHANK2*, and *ANKRD11*. The 22 genes significant in the missense category were *CCDC168, MECP2, PTEN, SCN2A, MUC19, PIEZO1, TBL1XR1, ZNF469, EML6, GREB1L, SLC6A1, OTOG, KDM5B, R3HCC1, CACNA1I, ACTG1, DDX3X, KCTD1, FREM3, KIR2DL3, LCORL*, and *PPP2R5D*. The 33 genes significant in the combined LOF and missense category were *SCN2A, CCDC168, PTEN, MECP2, ADNP, CHD8, FOXP1, KDM5B, CHD2, SHANK3, GRIN2B, MUC19, DDX3X, PIEZO1, MIB1, ZNF469, TBL1XR1, SLC6A1, SMAD6, OTOG, TLK2, GREB1L, EML6, POGZ, GIGYF1, R3HCC1, FREM3, PPP2R5D, BRSK2, CACNA1I, ACTG1, DNMT3A*, and *KCTD1*. Several of these genes have come up in other studies of autism (e.g., *SCN2A* ^31^, *CHD8* ^32^).

Next, we performed the same analyses in males with autism (n = 22,081 individuals) (**Figure 4A, Figure 4B, Figure 4C**). We list the significant genes in each category and label in parentheses after the gene as follows (**NSAF**) if there was not even nominal significance in females with autism for the same category. The 16 genes significant in the LOF category were *CHD8, SCN2A, SHANK3, FOXP1* (**NSAF**), *SMAD6* (**NSAF**), *PTEN, SYNGAP1, CHD2, MARK2, GIGYF1, ADNP, AUTS2* (**NSAF**), *KDM6B, MIB1, POGZ*, and *CCDC168* (**NSAF**). The 10 genes significant in the missense category were *CCDC168, PIEZO1* (**NSAF**), *MUC19, OTOG, EML6* (**NSAF**), *ZNF469, PTEN, ZNF84* (**NSAF**), *SCN2A*, and *IGSF23* (**NSAF**). The 19 genes significant in the combined LOF and missense category were *CCDC168, SCN2A, PTEN, FOXP1, CHD8, MUC19, PIEZO1* (**NSAF**), *CHD2, SMAD6* (**NSAF**), *KDM5B, ZNF469, OTOG* (**NSAF**), *ADNP, EML6* (**NSAF**), *SLC6A1* (**NSAF**), *GIGYF1* (**NSAF**), *SHANK3, FREM3* (**NSAF**), and *MIB1*. Overall, there were 12 unique genes that met exome-wide significant enrichment in males that were not nominally significant for the *same variant class* in females. These included *FOXP1, SMAD6, AUTS2, CCDC168, PIEZO1, EML6, ZNF84, IGSF23, OTOG, SLC6A1, GIGYF1*, and *FREM3*.

**Figure 4.**
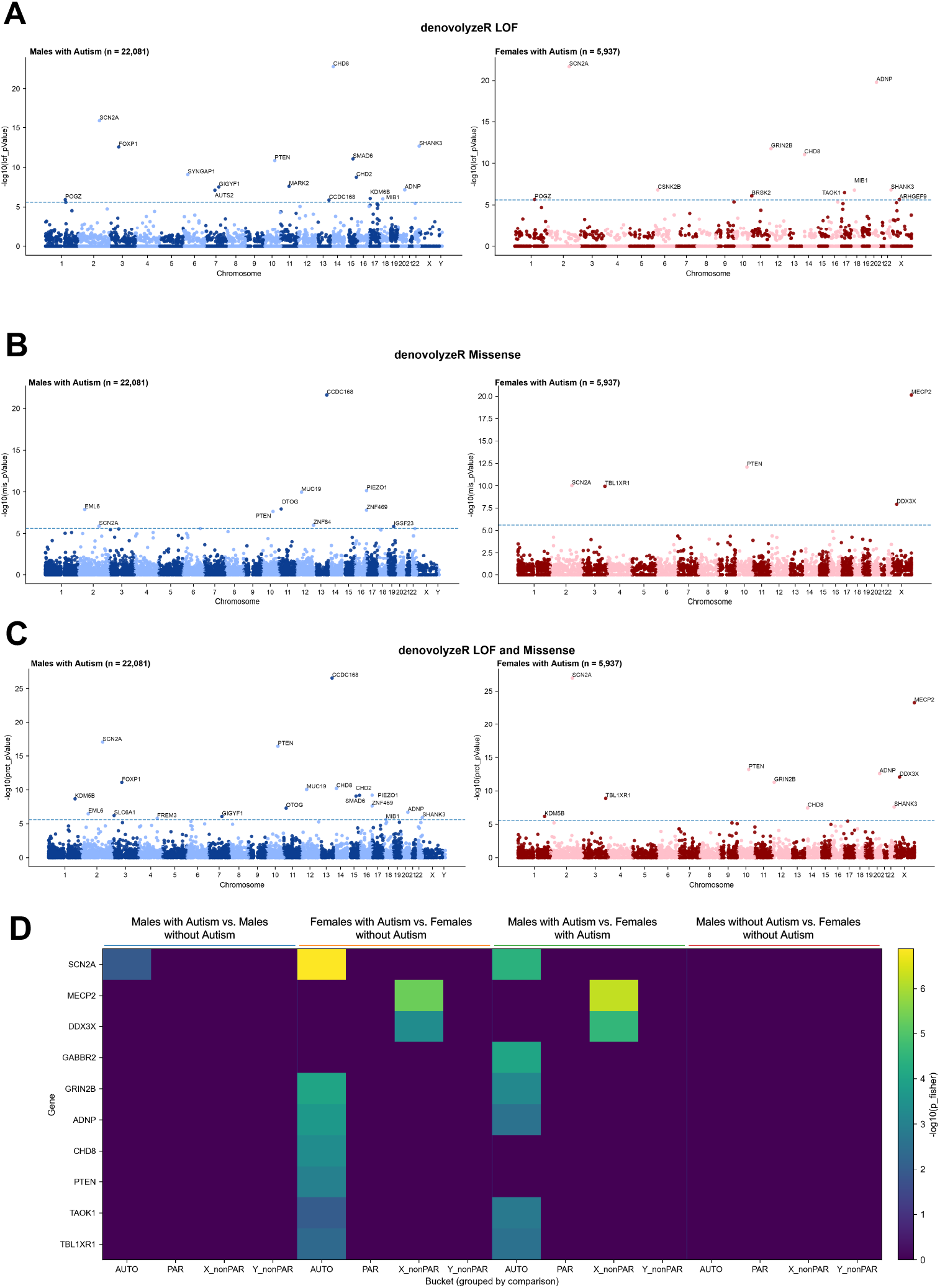
Gene-Based Enrichment Analyses. Shown in A) to C) are gene set enrichments analyses generated using denovolyzeR for two different settings (males with autism, females with autism) and shown in Figure D) are results from Fisher’s exact tests for four different comparison settings. A) Manhattan plots of denovolyzeR results for LOF DNVs, B) Manhattan plots of denovolyzeR results for missense DNVs, and C) Manhattan plots of denovolyzeR results for combined LOF and missense DNVs. D) Fisher’s exact test results with heatmap colors corresponding to the -log10(p-value). for the model of 0.45

Next, we performed the same analyses in females with autism (n = 5,937 individuals) (**Figure 4A, Figure 4B, Figure 4C**). We list the significant genes in each category and label in parentheses after the gene as follows (**NSAM**) if there was not even nominal significance in males with autism for the same category. The 11 genes significant in the LOF category were *SCN2A, ADNP, GRIN2B, CHD8, SHANK3, CSNK2B, MIB1, TAOK1* (**NSAM**), *BRSK2, POGZ*, and *ARHGEF9*. The 5 genes significant in the missense category were *MECP2* (**NSAM**), *PTEN, SCN2A, TBL1XR1* (**NSAM**), and *DDX3X* (**NSAM**). The 10 genes significant in the combined LOF and missense category were *SCN2A, MECP2* (**NSAM**), *PTEN, ADNP, DDX3X* (**NSAM**), *GRIN2B, TBL1XR1, SHANK3, CHD8*, and *KDM5B*. Overall, there were four genes that met exome-wide significant enrichment in females that were not nominally significant for the *same variant class* in males. These included *TAOK1, MECP2, DDX3X*, and *TBL1XR1*.

Next, we performed the same analyses in unaffected males (n = 6,481 individuals). No genes were significant in the LOF category. The one gene significant in the missense and the combined LOF and missense category was *STARD9*. Next, we performed the same analyses in unaffected females with autism (n = 6,868 individuals). No genes were significant in the LOF category. The 4 genes significant in the missense category were *MUC19, HEATR5A, WDFY4*, and *DNAH14*. The 3 genes significant in the combined LOF and missense category were *MUC19, HEATR5A*, and *WDFY4*.

Finally, we performed Fisher’s exact test analyses in four different comparison groups including males with autism versus males without autism, females with autism versus females without autism, males with autism versus females with autism, and males without autism versus females without autism (**Figure 4D**). Only one gene was exome-wide significant and that was *SCN2A* in the females with autism versus females without autism. Though not significant, other genes met the expected trends including *MECP2* and *DDX3X* trending towards enrichment in females with autism, specifically. One intriguing gene was *GABBR2* that trended towards significance in females in only the comparison of males with autism versus females with autism.

### Promoter-Based Testing of DNVs Reveals Enrichment of DNVs in RNU2-2

We evaluated all RefSeq gene promoters, in the WGS data only, across four groups: males with autism, females with autism, unaffected males, and unaffected females. A single promoter region, for two separate transcripts of *WDR74* (chr11:62840210-62842210 and chr11:62840156-62842156) reached genome-wide significance for DNVs (p = 3.8×10^-13^), and this signal was specific to males with autism (**Figure 5A**). Follow-up analyses revealed that most DNVs within this promoter clustered in the noncoding RNA gene *RNU2-2*, which overlaps a subregion of the promoter (**Figure 5B**). We therefore tested *RNU2-2* directly and observed a highly significant enrichment (p < 2.2 × 10^-16^) in males with autism (**Figure 5C**). We further looked for DNVs throughout the other data and found that DNVs in *RNU2-2* are present in 0.19% of males with autism and 0.20% of females with autism (**Figure 5D**). They are also present in 0.10% of unaffected males and were not observed in any unaffected females (**Figure 5E**). Secondary structures of wild type (**Figure 6A**) and variant haplotypes were generated with some variant haplotypes examining DNVs with or without inherited variants that highly destabilized the folding of the RNA (example in **Figure 6B**). By *in silico* saturation mutagenesis and folding-based metrics we identified the consequences of single base changes throughout the RNA (**Figure 6C**) and again showed that within the realm of all possible changes some variants present in individuals with autism in this study were predicted to be highly destabilizing (**Figure 6D**).

**Figure 5.**
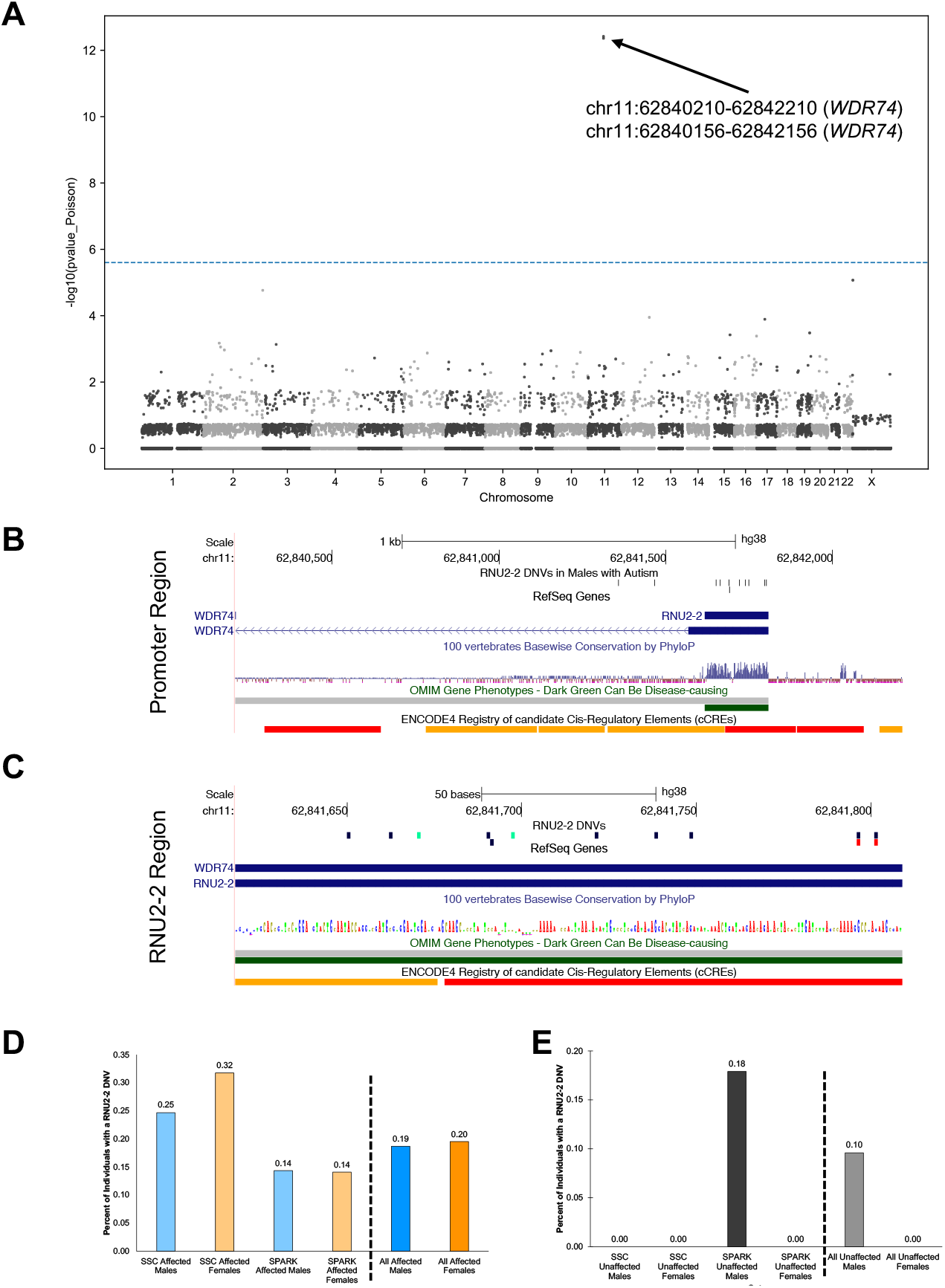
Promoter-Based Testing Reveals Enrichment of DNVs in the *RNU2-2* gene, which resides in the promoter of *WDR74*. A) Manhattan plot of -log10(p-value) versus chromosome in males with autism. Each point represents a promoter. The top promoter is genome-wide significant and is a promoter of the *WDR74* gene. B) Careful examination of DNVs in the promoter reveals localization of them to the *RNU2-2* noncoding RNA gene within the promoter region. C) Shown are the DNVs, in *RNU2-2*, in males with autism (navy), females with autism (red), and males without autism (green). Two of the DNVs are recurrent in unrelated individuals with autism. D) Percent of individuals with autism that contain a DNV in *RNU2-2* in each individual cohort of WGS data and in the combined set. E) Percent of individuals without autism that contain a DNV in *RNU2-2* in each individual cohort of WGS data and in the combined WGS data.

**Figure 6.**
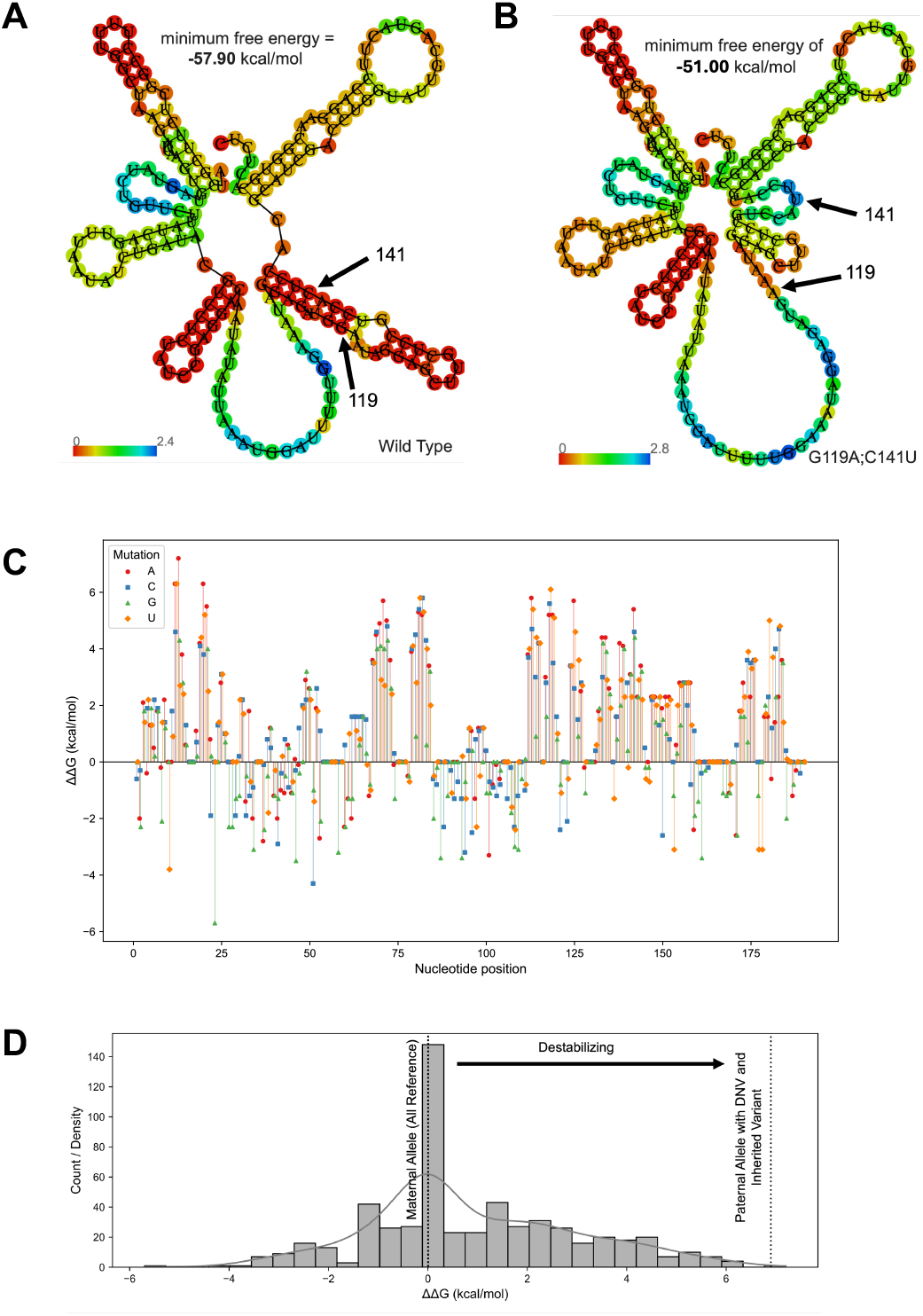
*In Silico* Assessment of Variants in *RNU2-2*. For A) and B) the positional entropy is displayed on the centroid secondary structure. A) Shown is the computational prediction of folding for the wild type version of the RNA as predicted by RNAfold. B) Computational prediction for folding of the RNA as present in an individual with autism. This individual contains both an inherited and a DNV on the paternal chromosome of origin so the structure is modeled in the context of both variants (G119A and C141U). C) Saturation mutagenesis screen of folding differences between every possible single change in the RNA and the wild type version of the RNA. D) Density and histogram plot showing the distribution of scores from C) and where the individual with autism shown in B) scores in terms of their maternal and paternal (contains the DNV) haplotypes. This individual has predicted destabilization of the RNA folding on the paternal allele.

## DISCUSSION

In this study, we investigated DNVs in autism, with a particular focus on differences by sex and diagnostic status. Whereas other studies of sex bias in autism have included individuals with neurodevelopmental disorders more broadly, this study focuses on the primary autism phenotype. Using data from three autism family cohorts, we generated high-quality DNV callsets for individuals with and without autism. These callsets enabled a range of complementary analyses, including: (1) testing enrichment across variant and gene-class categories; (2) identifying genes reaching exome-wide significance in males with autism, females with autism, males without autism, and females without autism; (3) directly comparing DNV enrichment between males and females across multiple comparison groups; and (4) evaluating the enrichment of noncoding variants. Importantly, we performed analyses that were aware of sex chromosomes including PARs.

The foundation of this study is in the generation of a high-quality DNV callset. This was done by developing a sex-aware DNV caller and applying stringent criteria based on multiple variant-level characteristics to get to the final DNVs. Rather than filtering DNVs using genomic annotations (e.g., repeat regions), we used intensive, feature-based interrogation of each candidate DNV to produce a robust, high-confidence callset. This opens up new regions of the genome to investigation in the context of autism. After generating the high-confidence DNV callset, we tested enri chment across predefined gene sets stratified by DNV class. As expected, synonymous DNVs showed no significant gene-set enrichments. Consistent with prior observations, we observed enrichment of missense and LOF DNVs in individuals with autism. Additional enrichments were detected in females, including gene sets on the non-PAR regions of the X chromosome and genes with sex-biased expression in the brain. Several gene sets are relatively small (e.g., hormone receptor genes); although they showed large enrichment, they did not remain significant after multiple-testing correction. These signals may warrant followup in larger cohorts.

Analyses of gene-specific enrichments by sex and diagnostic category, were insightful. Few genes met enrichment in any category in individuals *without* autism as one would expect. Several genes were enriched in individuals with autism and by performing sex-stratified analyses we could pinpoint genes that were enriched only in one sex for the DNV class being tested. These genes included genes previously implicated in autism and some newly implicated genes. Those specific to males included *FOXP1, SMAD6, AUTS2, CCDC168, PIEZO1, EML6, ZNF84, IGSF23, OTOG, SLC6A1, GIGYF1*, and *FREM3* and to females included *TAOK1, MECP2, DDX3X*, and *TBL1XR1*. Both *MECP2* and *DDX3X* are well-established genes with known enrichment in females. Some genes of interest from these lists include the newly implicated *PIEZO1*, which is a mechanosensitive ion channel and is important in the brain ^33, 34^ and *TAOK1*, which is involved in a neurodevelopmental disorder ^35^ wherein it has recently been reported that endocrinopathies are also common ^36^. One intriguing gene in the comparison of males with autism to females with autism was the gene *GABBR2* with enrichment of DNVs in females. While not exome-wide significant, this is a gene that may be of interest in future studies as it is the only one that was not an X chromosome gene that was close to significance. The neurodevelopmental disorder associated with *GABBR2* variants resembles Rett syndrome^37^ (typically caused by variants in the X-linked gene *MECP2*). Additionally, *GABBR2* variants have been reported in association with Rett-like phenotypes in individuals with classical Rett syndrome caused by *MECP2* variants ^*38*^.

In the final part of this study, we focused on promoters as they have previously been implicated in bulk as important for autism ^39, 40^. Herein, we queried each promoter for significant excess of DNVs and identified the *WDR74* promoter as significant specifically in males with autism. By inspection of the promoter in the UCSC Genome Browser^28^, we found that the *RNU2-2* noncoding RNA gene was located within this promoter and nearly all the DNVs were within it. This led us to hypothesize that the signal may be from this gene, and we found that *RNU2-2* was also genome-wide significant for excess DNVs. *RNU2-2* DNVs are present in 0.2% of males with autism. We further found that DNVs occur in females with autism as well and the overall rate is at 0.2% of individuals with autism. There were two males without autism that also had a DNV leading to an overall rate of 0.1% in unaffected males. Assessment of females without autism identified no DNVs in this region. These results indicate that *RNU2-2* may be an important gene related to autism in addition to the severe neurodevelopmental disorder for which it is already associated with in the literature ^41^. Investigation of variants identified in individuals with autism in relationship to RNA folding, *in silico*, identified potential destabilization of the RNA in some individuals with autism. Further assessment of this significant finding is imperative.

## Data Availability

Approved researchers can obtain the SSC population dataset described in this study by applying at https://base.sfari.org. Approved researchers can obtain the SPARK population dataset described in this study by applying at https://base.sfari.org.

## Acknowledgements

We are grateful to all of the families at the participating Simons Simplex Collection (SSC) sites, as well as the principal investigators (A. Beaudet, R. Bernier, J. Constantino, E. Cook, E. Fombonne, D. Geschwind, R. Goin-Kochel, E. Hanson, D. Grice, A. Klin, D. Ledbetter, C. Lord, C. Martin, D. Martin, R. Maxim, J. Miles, O. Ousley, K. Pelphrey, B. Peterson, J. Piggot, C. Saulnier, M. State, W. Stone, J. Sutcliffe, C. Walsh, Z. Warren, E. Wijsman). We appreciate obtaining access to SSC whole-genome sequencing data on SFARI Base. Approved researchers can obtain the SSC population dataset described in this study by applying at https://base.sfari.org. We are grateful to all the families in SPARK, the SPARK clinical sites and SPARK staff. We appreciate obtaining access to SPARK whole-exome sequencing and whole-genome sequencing data on SFARI Base. Approved researchers can obtain the SPARK population dataset described in this study by applying at https://base.sfari.org.

